# Corticosteroid Treatment and Hemorrhage Risk in Cerebral Amyloid Angiopathy with Cortical Superficial Siderosis: A Matched Cohort Analysis

**DOI:** 10.1101/2025.09.24.25336535

**Authors:** Philip Arndt, Eya Khadhraoui, Sebastian J. Müller, Katja Neumann, Hendrik Mattern, Sarah-Marie Löding, Melis Tas, Sven G. Meuth, Valentina Perosa, Andreas Charidimou, Stefanie Schreiber

## Abstract

Cortical superficial siderosis (cSS) is the strongest marker of future intracranial hemorrhage (ICH) in cerebral amyloid angiopathy (CAA), a condition without disease-modifying therapy. Emerging evidence suggests that cSS may reflect meningovascular inflammation potentially amenable to immunosuppression.

We conducted a single-center matched cohort study comparing high-dose corticosteroid therapy versus no immunosuppression in patients with probable CAA and cSS. Treated patients were matched 1:2 to untreated patients by demographics, cSS multifocality, and baseline intracerebral hemorrhage. The primary outcome was incident ICH; secondary outcomes were any cerebrovascular event and mortality. Kaplan–Meier analysis assessed event-free survival. cSS progression was evaluated in patients with ≥6 months MRI follow-up, and inflammation in a subgroup with post-contrast vessel-wall-imaging (VWI).

Thirty-six patients were included (12 treated, 24 untreated; mean age 77.4 years; 91% disseminated cSS). Over a median follow-up of 2.23 years, 11 patients had cerebrovascular events (15 ICHs, 3 ischemic strokes), all in the untreated group (p=0.041 for ICH; p=0.037 for any event). Mortality did not differ (p=0.634). cSS progressed in 2/6 (33%) treated versus 4/7 (57%) untreated patients. Baseline VWI detected inflammation in 7/9 patients (78%), which regressed after corticosteroids.

Corticosteroid therapy was associated with fewer cerebrovascular events in high-risk CAA, supporting prospective controlled validation.

## Background

Sporadic cerebral amyloid angiopathy (CAA) is a common age-associated cerebral small vessel disease and a major cause of intracranial hemorrhage (ICH), including lobar intracerebral hemorrhage, convexity subarachnoid hemorrhage (SAH) and non-traumatic subdural hematoma (SDH).^1,2^ Among its putative neuroimaging biomarkers, cortical superficial siderosis (cSS) - reflecting hemosiderin deposition along the cortical surface from chronic leptomeningeal bleeding – has emerged as the strongest independent predictor of future hemorrhage risk.^3^

In a subset of patients, CAA is accompanied by an autoinflammatory response, known as CAA-related inflammation (CAA-ri), characterized by vasogenic edema and often responsive to corticosteroid therapy.^4^ However, emerging evidence suggests that even in the absence of typical or overt CAA-ri, cSS may reflect localized meningovascular inflammation.^5-8^ This raises the possibility that immunosuppressive therapy could modify disease course in CAA patients with disseminated cSS, even outside the classical CAA-ri syndrome. We therefore conducted a matched cohort study to examine whether corticosteroid treatment is associated with reduced risk of cerebrovascular events in patients with probable CAA and cSS.

## Methods

### Study population and baseline data collection

We included consecutive patients enrolled between March 2007 and April 2025 in our prospectively maintained CAA registry at the Department of Neurology, Otto-von-Guericke University Magdeburg. Inclusion criteria were: (1) a diagnosis of probable CAA according to the Boston criteria v2.0; (2) presence of cSS on brain MRI; and (3) available follow-up data on cerebrovascular events and mortality. Patients who received corticosteroids were identified from the registry and matched 1:2 with untreated patients using manual nearest-neighbor matching without replacement. Matching variables included age, sex, cSS multifocality (disseminated vs. focal), and presence of baseline intracerebral hemorrhage. Investigators performing the matching were blinded to outcomes. Matching adequacy was evaluated through between-group comparisons of baseline characteristics.

### Outcome analysis

The primary outcome was incident ICH, including spontaneous lobar intracerebral hemorrhage, non-aneurysmal convexity SAH, and non-traumatic SDH. Secondary outcomes included (1) a composite of any cerebrovascular event (ICH or ischemic stroke), and (2) all-cause mortality. The baseline timepoint was defined as the MRI at corticosteroid initiation (treatment group) or the first MRI demonstrating cSS (control group). Follow-up data were obtained from our prospective registry, electronic medical records review, radiology databases, and structured interviews with patients or caregivers.

### MRI acquisition and analysis

MRI scans were acquired using clinical 3T (n=17) or 1.5T (n=15) scanners (Siemens Healthineers, Erlangen, Germany). All MRI studies included T2^*^-weighted gradient-recalled echo (GRE) sequences and were evaluated by two senior neuroradiologists (E.K. and S.M.) according to STRIVE criteria and MARS.^9,10^ cSS was defined as curvilinear hypointense signal along the cortical surface according to consensus guidelines.^11^ Each hemisphere was rated for cSS multifocality (0–2 points), yielding a total score of 0–4. Disseminated cSS was defined as >3 adjacent sulci or ≥2 nonadjacent foci.^12^ For patients with follow-up imaging ≥6 months apart, baseline and follow-up T2^*^-GRE scans were visually assessed and compared side-by-side for cSS progression, defined as one or more new foci not present at baseline.^13^

For some patients high-resolution pre- and post-contrast vessel-wall imaging (VWI) was available. It was acquired after administration of Gadovist using a transversal, single-slab, non-selective 3D turbo spin echo sequence with generalized autocalibration partial parallel acquisition for image acceleration and spectral attenuated inversion recovery for fat suppression. Whole-brain coverage was achieved with a 200 mm field of view, isotropic voxel size of 0.8 mm, repetition time of 700 ms, and echo time of 23 ms. All scans were performed on a 3-Tesla MRI system (Philips Achieva, Best, Netherlands). VWI was assessed separately for large vessels (A1–A3, M1–M3, P1–P3, vertebral arteries, basilar artery), medium-sized leptomeningeal vessels, and small cortical vessels. The number of contrast-enhancing vessel walls was counted. All enhancement findings were cross-checked with pre-contrast VWI sequences to minimize artifacts. If uncertainty remained regarding whether an enhancing structure was arterial, it was not classified as vessel wall enhancement. Veins were differentiated based on their typical anatomical trajectory, continuity, and morphology.

### Protocol Approvals

This study analyzed data from patients treated with corticosteroids as part of an individualized therapeutic approach in the absence of established alternatives. All patients were fully informed about the off-label use of corticosteroids, including potential risks, and provided written informed consent. The treatment approach and data use were reviewed by the local ethics board, which confirmed that publication of anonymized patient data was permissible under these conditions. The study was conducted in accordance with institutional and national regulations.

### Data Availability

Data are available from the corresponding author upon reasonable request.

### Statistical analysis

Continuous variables were reported as mean (SD) or median (IQR) and categorical variables as proportions. Group comparisons used t-tests, Mann–Whitney U, χ^2^ or Fisher’s exact tests as appropriate. Kaplan–Meier survival analysis with log-rank testing was used to assess the association between corticosteroid treatment and time to outcome events. Time-to-event was calculated from baseline MRI to the first qualifying event or censoring. A two-sided p-value <0.05 was considered significant. Analyses were performed using IBM SPSS Statistics 28.0 and GraphPad Prism 10.0.

## Results

Among 36 patients with probable CAA and cSS, 12 received corticosteroid therapy and were matched to 24 untreated controls. The mean age was 77.4 years (SD 7.3), 47% were female, and 33% initially presented with lobar intracerebral hemorrhage. cSS was disseminated in 92% and focal in 8% of patients. Baseline clinical and imaging characteristics were similar between groups (**Table 1**). Follow-up duration was significantly longer in the untreated group (median 2.86 years, IQR 1.50–7.83) compared to the corticosteroid group (median 0.79 years, IQR 0.29–3.34; p=0.045), reflecting the more recent and systemic adoption of corticosteroid therapy at our center. Prior to 2022, corticosteroids were used sporadically in patients with cSS. Beginning in 2022, following encouraging results from an individual treatment attempt,^8^ corticosteroid therapy was routinely offered to all eligible patients with cSS (1000 mg intravenous methylprednisolone for 3–5 days). No randomized treatment assignment was performed.

**Table 1.** Baseline characteristics of all CAA patients with cortical superficial siderosis, stratified by corticosteroid treatment status. Continuous variables are expressed as mean (standard deviation) or median (interquartile range) as appropriate; categorical variables are expressed as proportions. Abbreviations: cSS, cortical superficial siderosis; MB, microbleeds; TFNE, transient focal neurological episodes SD, standard deviation.

|  | All patients<br>with cSS<br>n = 36 | Patients<br>without any<br>treatment<br>n = 24 | Patients treated<br>with<br>corticosteroids<br>n = 12 | p-value |
| --- | --- | --- | --- | --- |
| **Baseline characteristics** |  |  |  |  |
| Mean age (SD) | 77.4 (7.3) | 77.9 (6.9) | 76.4 (8.2) | 0.540 |
| Female | 17 (47%) | 12 (50%) | 5 (42%) | 0.637 |
| Intracerebral hemorrhage | 11 (31%) | 7 (29%) | 4 (33%) | 1.000 |
| TFNE | 11 (31%) | 7 (29%) | 4 (33%) | 1.000 |
| Cognitive impairment | 12 (33%) | 8 (33%) | 4 (33%) | 1.000 |
| Seizure | 11 (31%) | 8 (33%) | 3 (25%) | 0.705 |
| Antiplatelet usage | 12 (33%) | 7 (29%) | 5 (42%) | 0.643 |
| Anticoagulant usage | 3 (8%) | 2 (8%) | 1 (8%) | 1.000 |
| History of myocardial infarct | 2 (6%) | 1 (5%) | 1 (8%) | 1.000 |
| Arterial hypertension | 29 (81%) | 19 (79%) | 10 (83%) | 0.610 |
| Type 2 diabetes | 3 (8%) | 1 (5%) | 2 (17%) | 0.538 |
| Dyslipidemia | 16 (44%) | 10 (42%) | 6 (50%) | 0.895 |
| **MRI markers** |  |  |  |  |
| Disseminated cSS | 33 (92%) | 22 (92%) | 11 (92%) | 1.000 |
| Median cSS score (IQR) | 2 (2-3) | 2 (2-3) | 3 (2-4) | 0.510 |
| cSS score 1 | 3 (9%) | 2 (8%) | 1 (8%) | 1.000 |
| cSS score 2 | 15 (42%) | 11 (46%) | 4 (33%) | 0.721 |
| cSS score 3 | 8 (22%) | 6 (25%) | 2 (17%) | 0.691 |
| cSS score 4 | 10 (28%) | 5 (21%) | 5 (42%) | 0.188 |
| Median lobar MB counts (IQR) | 5 (2-11) | 5 (3-10) | 5 (2-23) | 0.878 |

During a median follow-up of 2.23 years (124.60 patient-years), 11 patients (31%) experienced cerebrovascular events, including 15 ICH (11 lobar intracerebral hemorrhages, 3 convexal SAH, 1 SDH) and 3 ischemic strokes. Four patients had multiple events. Kaplan–Meier analysis showed that corticosteroid therapy was associated with longer event-free survival for both ICH (p=0.041) and the composite cerebrovascular event outcome (p=0.037). No cerebrovascular events occurred in the corticosteroid treated group. This corresponded to an ICH rate of 15.2 and an ischemic stroke rate of 3.0 per 100 patient-years in the untreated group, versus 0.0 per 100 patient-years in the treated group. All-cause mortality occurred in 14 patients (58%) in the untreated group and in 3 patients (25%) in the corticosteroid group (log-rank p=0.634).

Follow-up T2^*^-GRE MRI data ≥6 months apart were available in 13 patients (6 treated, 7 untreated). cSS progression occurred in 2 of 6 treated patients (33%; median interval: 1.4 years; range: 0.6–4.7), and 4 of 7 untreated patients (57%; median interval: 1.2 years; range: 0.8–7.1).

Contrast-enhanced VWI was available for nine patients. The majority of patients showed neuroimaging evidence of inflammation on VWI, including leptomeningeal enhancement (n=7 patients, 78%) or cortical vessel wall enhancement (n=6 patients, 67%). Vessel wall enhancement was pronounced in areas affected by cortical superficial siderosis (cSS), but was also observed distant from cSS. Five patients exhibited diffuse vessel wall enhancement throughout the brain (**Table 2, Figure 2**). Enhancement of large-caliber arteries was seen in three patients (33%). In three patients treated with high-dose corticosteroids, short term follow-up VWI demonstrated a responsive regression of vessel wall enhancement.

**Table 2.** MRI markers of inflammation on post-contrast vessel wall imaging.

| Case | Cortical superficial siderosis | Sulcal HI on native FLAIR | LME | Counts of LM VWE | Counts of cortical VWE | Large VWE | Treatment for inflammation |
| --- | --- | --- | --- | --- | --- | --- | --- |
| 1 | bilateral, disseminated | yes | bilateral, diffuse | >100 | 4 | right M3 | cortico-steroids for 3d |
| 2 | right, disseminated | yes | bilateral, diffuse | >100 | 8 | no | no |
| 3 | bilateral, disseminated | yes | bilateral, diffuse | >100 | 3 | no | cortico-steroids for 3d |
| 4 | left, focal | yes | bilateral, diffuse | 71 | 7 | no | cortico-steroids for 5d |
| 5 | bilateral, disseminated | yes | bilateral, parieto-temporal | 41 | 0 | no | cortico-steroids for 3d |
| 6 | bilateral, disseminated | yes | bilateral, diffuse | 28 | 3 | Bilateral M2 and M3 | cortico-steroids for 3d |
| 7 | bilateral, disseminated | yes | bilateral, fronto-parietal | 16 | 5 | left M1, right M2 | cortico-steroids for 3d |
| 8 | left, focal | no | No | 0 | 0 | no | cortico-steroids for 3d |
| 9 | right, disseminated | no | No | 0 | 0 | no | cortico-steroids for 3d |

Abbreviations: d, days; HI, hyperintensities; LM, leptomeningeal, MRI, magnetic resonance imaging; VWE, vessel wall enhancement

**Figure 1.**
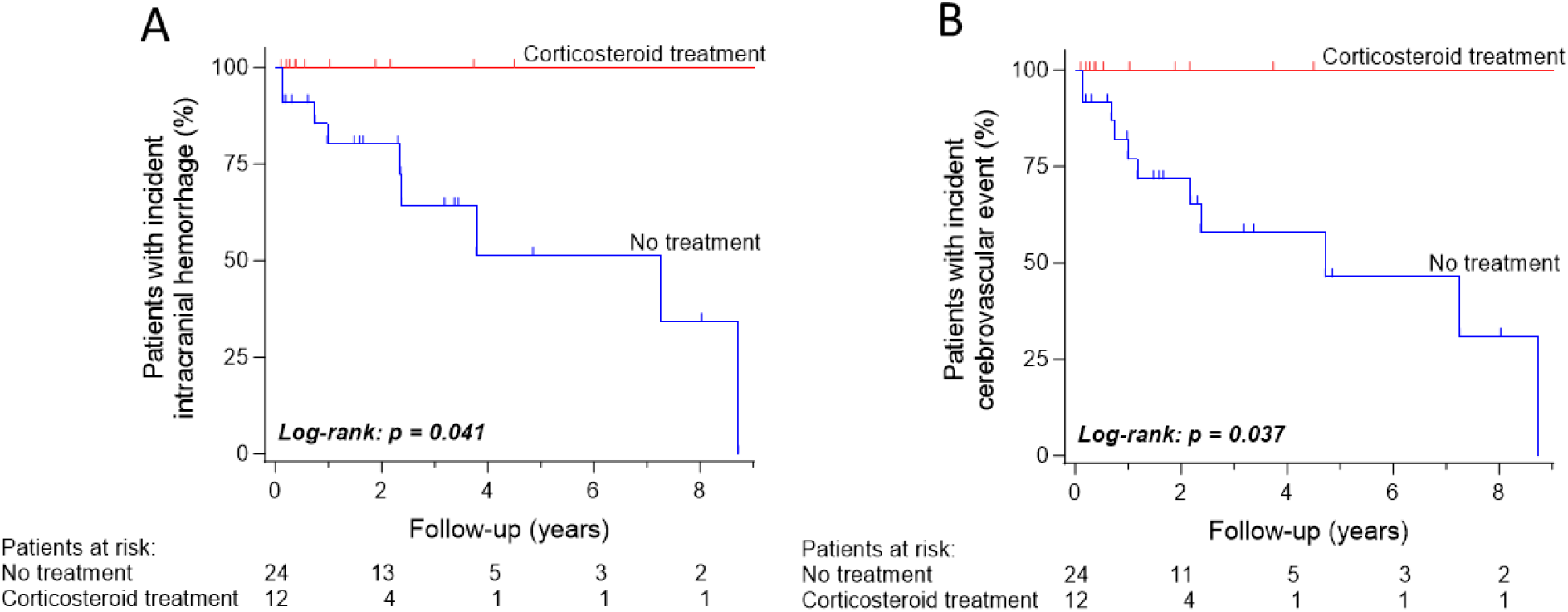
Kaplan–Meier survival curves stratified by corticosteroid treatment. (A) Time to first incident intracranial hemorrhage, including lobar intraparenchymal hemorrhage, non-aneurysmal convexity subarachnoid hemorrhage, and non-traumatic subdural hemorrhage. (B) Time to first cerebrovascular event (composite of hemorrhage and ischemic stroke). Tick marks indicate censored observations.

**Figure 2.**
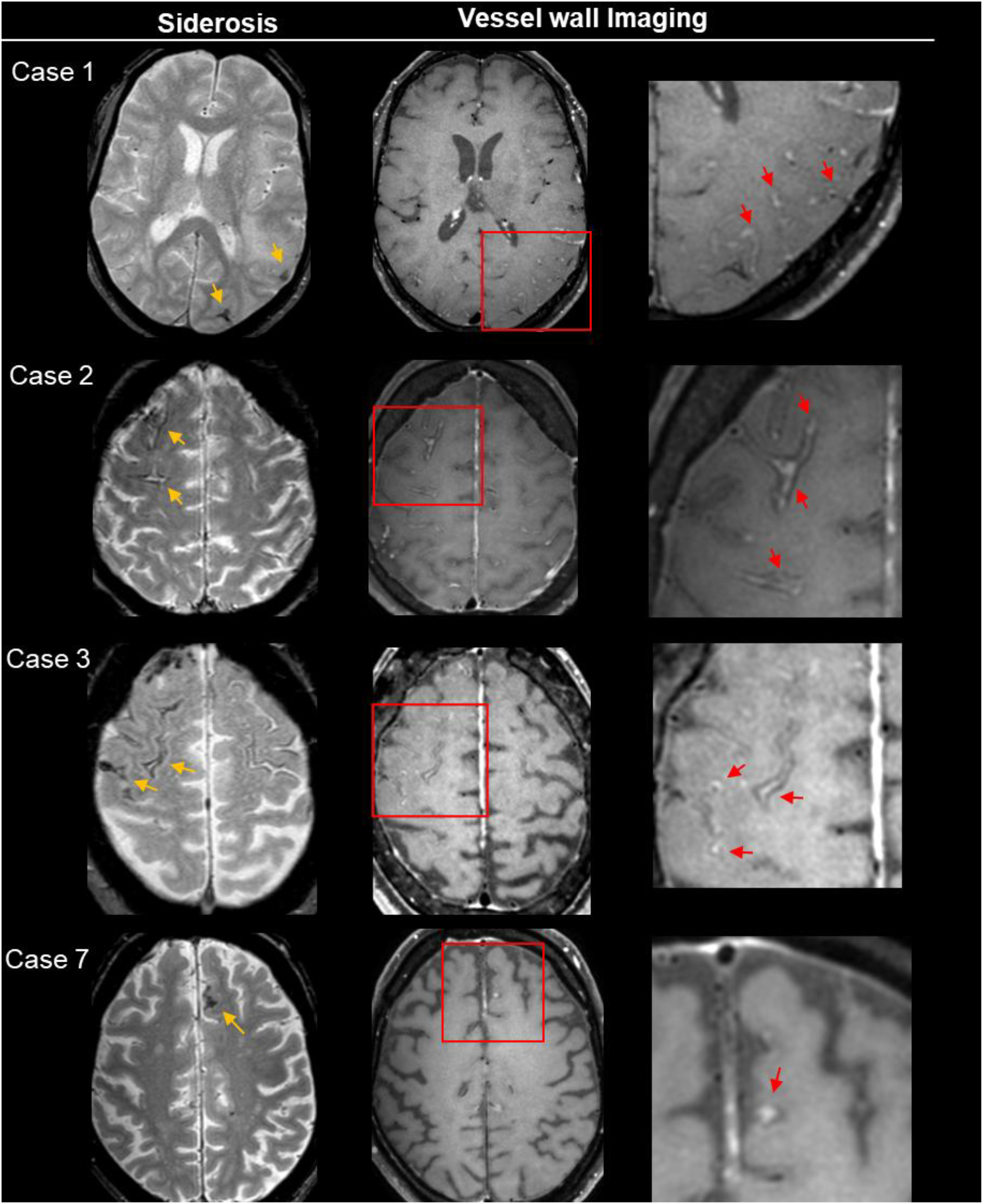
Anatomical overlap of cortical superficial siderosis and vessel wall enhancement on brain MRI. Four representative cases illustrate the variability in the severity of vessel wall enhancement on post-contrast vessel wall imaging. Cases 1 - 3 demonstrate severe enhancement with typical “donut-signs”, and case 7 shows moderate enhancement. Corresponding imaging variables for each patient are demonstrated in **Table 2**. Image interpretation was guided by pre-contrast sequences and anatomical landmarks to avoid venous misclassification.

## Discussion

In this matched cohort study of patients with probable CAA and cSS, corticosteroid therapy was associated with complete absence of cerebrovascular events during follow-up, in contrast to a substantial event rate among untreated controls. While the findings are limited by small sample size and the non-randomized, observational design, the magnitude of this difference raises the possibility of a protective effect of corticosteroids on hemorrhagic risk in this high-risk population with advanced CAA.

To our knowledge, this is the first study to systematically evaluate cerebrovascular outcomes in probable CAA patients with cSS treated with corticosteroids outside the classical CAA-ri syndrome. While CAA-ri is well recognized as corticosteroid-responsive,^4^ emerging literature suggests that meningovascular inflammation may extend beyond this defined syndrome, including a wider spectrum of CAA presentations, such as multifocal cSS. This hypothesis is supported by vessel wall imaging demonstrating contrast enhancement adjacent to cSS foci in 78% of patients. This is in line with a recent case series of CAA patients with transient focal neurological deficits and cSS, in which 5 out of 6 (83%) patients showed vessel wall enhancement ^5,6^ Our findings build on these observations, suggesting that timely immunosuppression may favorably alter clinical outcomes in a broader subset of CAA patients.

Radiological follow-up in a small subgroup of patients showed cSS progression in 57% of untreated patients compared to 33% of those treated with corticosteroids. Although MRI follow-up intervals were heterogeneous and the sample size limited, these preliminary findings further support the hypothesis that corticosteroid therapy may attenuate disease progression, potentially by modulating inflammation-driven vessel wall injury. Our observations are consistent with prior data demonstrating cSS progression in 50% of patients with sporadic CAA and cSS over one year period.^13^

Recent proteomic evidence lends further support to the hypothesis of autoimmune mechanisms in sporadic CAA. Vascular proteomic analysis of brain autopsy tissue, revealed upregulation of complement cascade proteins - including C3, C4A, C1Q and APCS - alongside other regulators of the humoral immune response.^14^ Additionally, corticosteroids have been shown to mitigate the proinflammatory and cytotoxic effects of beta-amyloid in cultured human cerebrovascular smooth muscle cells.^15^ Taken together with our clinical findings, these observations strengthen the concept of an immune-responsive disease spectrum in some patients with advanced CAA beyond classical CAA-ri.

### Limitations

The small sample size and the observational non-randomized design limit generalizability and causal inference. Follow-up was significantly shorter in the corticosteroid-treated group, reflecting the recent adoption of this therapy, and may have underestimated event rates in this group. The absence of cerebrovascular events in the treatment group precluded multivariable adjustments. Furthermore, imaging protocols were non-standardized, introducing potential measurement variability. Despite these limitations, the consistency of findings across clinical and imaging outcomes, offers preliminary support for a potential therapeutic effect and underscores the need for prospective, controlled validation.

## Data Availability

All data produced in the present study are available upon reasonable request to the authors.

## Acknowledgments

This study was supported by the BB-DARS project, funded by the Deutsche Alzheimer Gesellschaft (DAlzG) and the Förderstiftung Dierichs.

## Author Contributions

S.S. contributed to the conception and design of the study; all authors contributed to the acquisition and analysis of data; P.A. and S.S. contributed to drafting the text; P.A. contributed to preparing the figures. All the authors contributed to a critical review of the manuscript.

